# Multi-Task Deep Learning for Predicting Metabolic Syndrome from Retinal Fundus Images in a Japanese Health Checkup Dataset

**DOI:** 10.1101/2025.05.13.25327551

**Authors:** Tohru Itoh, Koichi Nishitsuka, Yasufumi Fukuma, Satoshi Wada

**Author notes:** Corresponding author (KN). These authors contributed equally to this work (Tohru Itoh, Koichi Nishitsuka).

## Abstract

**Background:** Retinal fundus images provide a noninvasive window into systemic health, offering opportunities for early detection of metabolic disorders such as metabolic syndrome (METS).

**Objective:** This study aimed to develop a deep learning model to predict METS from fundus images obtained during routine health checkups, leveraging a multi-task learning approach.

**Methods:** We retrospectively analyzed 5,000 fundus images from Japanese health checkup participants. Convolutional neural network (CNN) models were trained to classify METS status, incorporating fundus-specific data augmentation strategies and auxiliary regression tasks targeting clinical parameters such as abdominal circumference (AC). Model performance was evaluated using validation accuracy, test accuracy, and the area under the receiver operating characteristic curve (AUC).

**Results:** Models employing fundus-specific augmentation demonstrated more stable convergence and superior validation accuracy compared to general-purpose augmentation. Incorporating AC as an auxiliary task further enhanced performance across architectures. The final ensemble model with test-time augmentation achieved a test accuracy of 0.696 and an AUC of 0.73178.

**Conclusion:** Combining multi-task learning, fundus-specific data augmentation, and ensemble prediction substantially improves deep learning-based METS classification from fundus images. This approach may offer a practical, noninvasive screening tool for metabolic syndrome in general health checkup settings.

## Introduction

Metabolic syndrome (METS) is a condition characterized by a combination of visceral fat accumulation and metabolic risk factors such as hypertension, dyslipidemia, and impaired glucose tolerance. It is known to significantly increase the risk of cardiovascular disease and type 2 diabetes [1,2]. As the global prevalence of METS continues to rise, early detection and risk stratification have become increasingly important in public health. Traditionally, diagnosing METS requires physical examination and blood tests, which may limit accessibility in resource-limited or non-clinical settings.

In recent years, advances in artificial intelligence (AI), particularly deep learning, have enabled the development of image-based diagnostic tools that can non-invasively estimate systemic health conditions from medical images [3,4]. Among such modalities, retinal fundus photography is especially promising, as the retinal vasculature and optic nerve can reflect systemic pathophysiological changes, positioning the retina as a “window to overall health” [5].

Poplin et al. demonstrated that deep learning could predict systemic risk factors such as blood pressure, gender, and smoking status from retinal images with high accuracy [6]. Furthermore, Cheung et al. showed that METS could be predicted from fundus images in over 13,000 subjects, achieving an area under the receiver operating characteristic curve (AUC) of 0.744 [7]. However, most existing models are developed using non-Japanese datasets, and few studies have explored METS prediction using fundus images in Japanese populations.

The Japan Ocular Imaging Registry (JOIR), a real-world ophthalmic image database, has provided access to high-quality retinal images from health checkup participants in Japan [8]. In this study, we aimed to develop a deep learning model for METS prediction using these fundus images and a limited set of associated clinical data. We employed a multi-task learning approach incorporating abdominal circumference (AC) as an auxiliary loss, implemented fundus-specific data augmentation strategies, and evaluated model robustness using ensemble learning and test-time augmentation (TTA). This approach demonstrated promising results and was recognized with the first-place award at an AI competition organized by the Japanese Society of Ophthalmic AI.

## Methods

### Dataset and Participants

The dataset used in this study was provided by the Japanese Society of Ophthalmic AI as part of an AI model development competition. It is based on the Japan Ocular Imaging Registry (JOIR) [8] and includes 5,000 anonymized retinal fundus photographs from Japanese male health checkup participants. All subjects were male, a standardization applied by the organizers to reduce variation during model training. An additional set of 500 cases was provided as an independent test dataset. Clinical variables accompanying the images included age, AC, systolic blood pressure (SBP), diastolic blood pressure (DBP), triglycerides (TG), high-density lipoprotein cholesterol (HDL), and fasting plasma glucose (FPG). The presence or absence of METS was annotated based on Japanese diagnostic criteria. This retrospective study used fully anonymized data obtained from routine health checkups conducted as part of a public health program. No identifying personal information was accessible to the authors. According to Japanese ethical guidelines and institutional policies, the use of such anonymized, non-interventional data does not require ethics committee approval.

### Image Preprocessing and Quality Control

Images with excessive blur, poor contrast, or pathological findings (e.g., diabetic retinopathy, macular scars, central retinal artery occlusion) were excluded. Additionally, cases with abdominal circumference values falling outside ±3 standard deviations were removed to mitigate the impact of outliers. After exclusion, 4,785 training cases and 500 test cases remained. Original fundus images were acquired at a resolution of 1920 × 1280 pixels. Each image was cropped without preserving the original aspect ratio and then resized according to the input requirements of each model architecture: 288 × 288 pixels for ConvNeXt-Base, and 256 × 256 pixels for SE-ResNeXt-50 and Swin Transformer V2 Base. All images were subsequently normalized prior to model input.

### Data Augmentation Strategies

To improve generalization under limited data conditions, we compared two types of image augmentation strategies during model training. The first strategy, referred to as Case 1, was specifically designed for retinal fundus images. It included anatomically conservative transformations such as small-angle rotation, brightness and contrast adjustment, color saturation modulation, and local contrast enhancement using CLAHE. Horizontal flipping was deliberately excluded in Case 1 to preserve the orientation of anatomical landmarks such as the optic disc and macula. This strategy aimed to maintain biological plausibility while introducing sufficient variability to prevent overfitting.

The second strategy, Case 2, employed general-purpose augmentations commonly used in standard image classification tasks. This included transformations such as horizontal flipping, affine transformation, Gaussian noise injection, motion blur, and channel-wise color shifting. Unlike Case 1, Case 2 did not account for the unique anatomical features of retinal images, and therefore served as a baseline for comparison. The specific augmentation techniques included in each strategy are summarized in Table 1. The impact of each augmentation strategy on model performance was evaluated using five-fold cross-validation on the training dataset. Performance metrics reported in this study, however, are based solely on the independent 500-case test dataset.

**Table 1.**
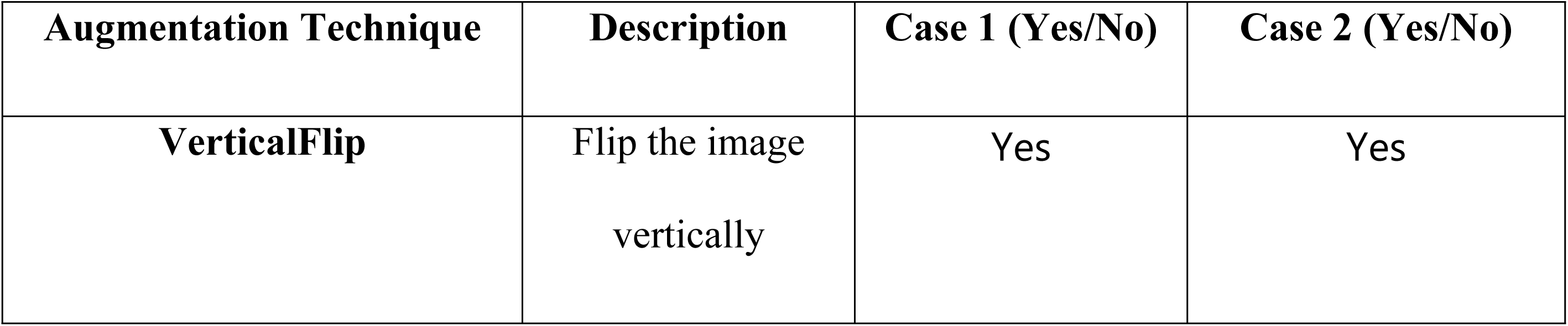
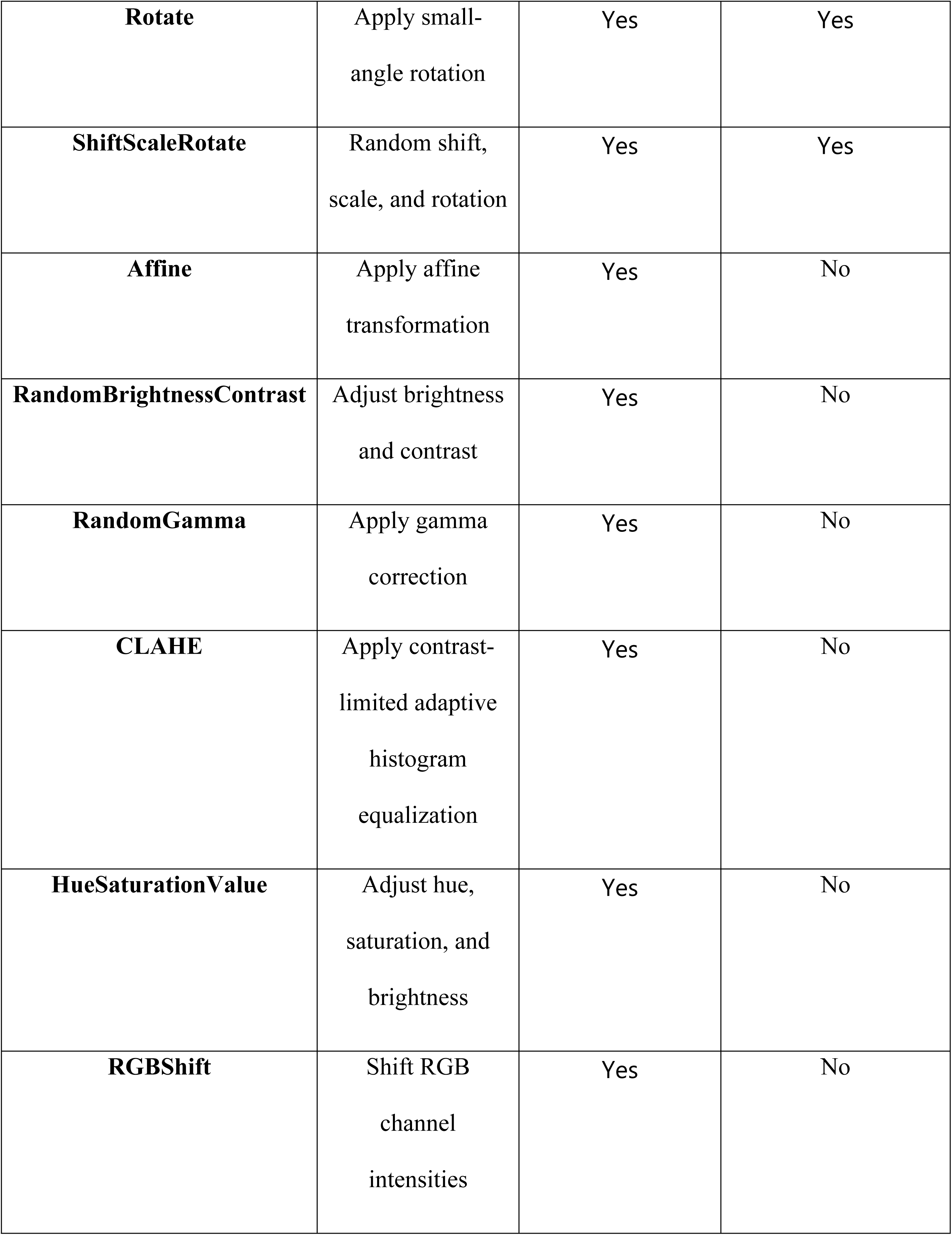
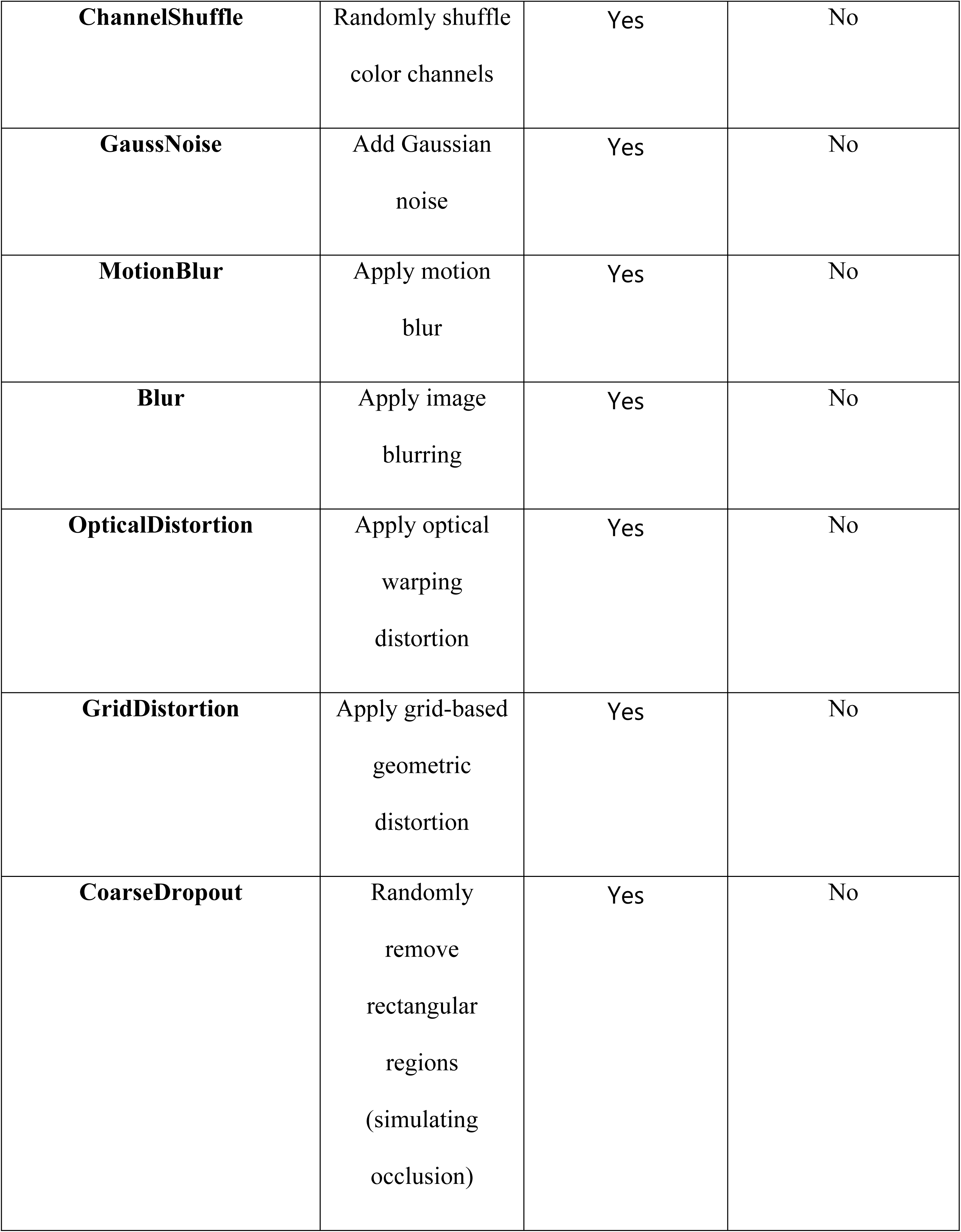
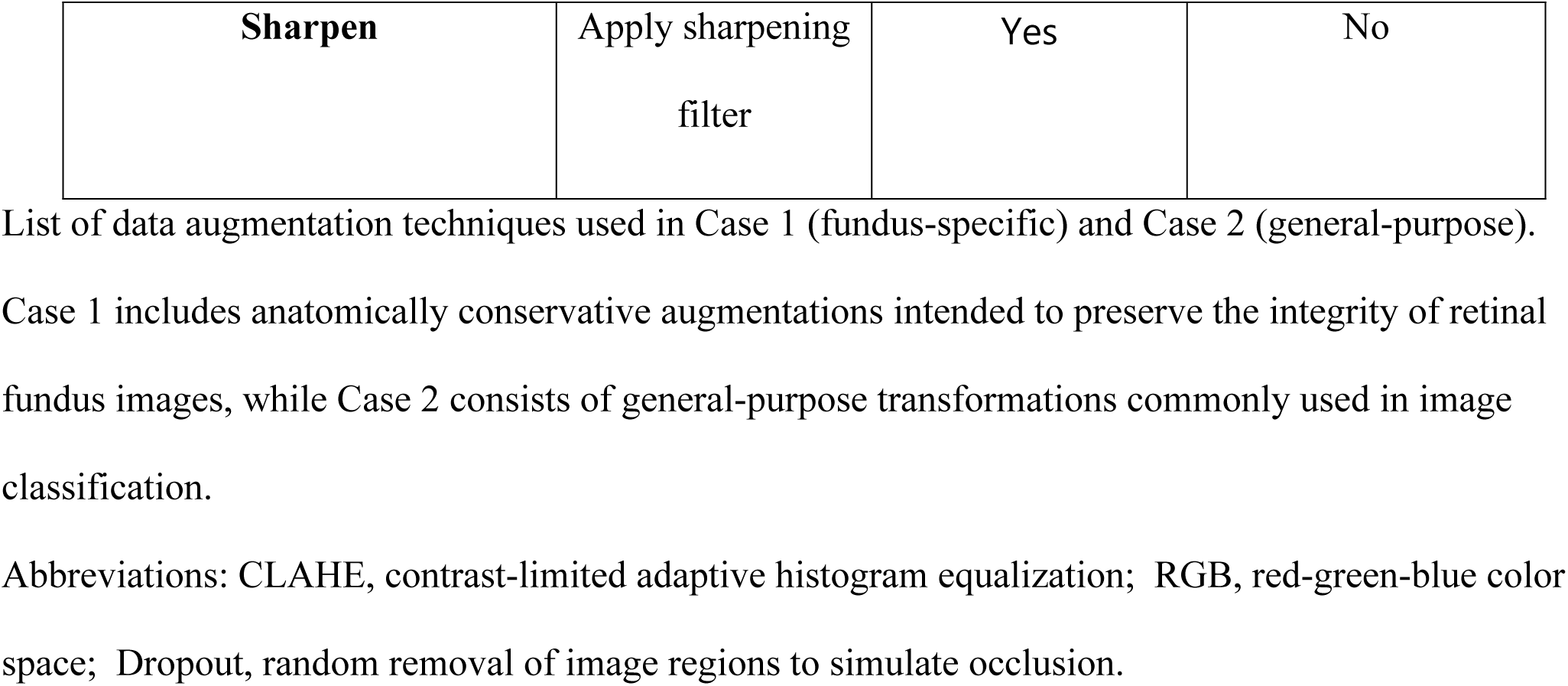
Comparison of data augmentation techniques used in Case 1 (fundus-specific) and Case 2 (general-purpose)

| Augmentation Technique | Description | Case 1 (Yes/No) | Case 2 (Yes/No) |
| --- | --- | --- | --- |
| VerticalFlip | Flip the image<br>vertically | Yes | Yes |
| **Rotate** | Apply small-angle rotation | Yes | Yes |
| **ShiftScaleRotate** | Random shift, scale, and rotation | Yes | Yes |
| **Affine** | Apply affine transformation | Yes | No |
| **RandomBrightnessContrast** | Adjust brightness and contrast | Yes | No |
| **RandomGamma** | Apply gamma correction | Yes | No |
| **CLAHE** | Apply contrast-limited adaptive histogram equalization | Yes | No |
| **HueSaturationValue** | Adjust hue, saturation, and brightness | Yes | No |
| **RGBShift** | Shift RGB channel intensities | Yes | No |
| **ChannelShuffle** | Randomly shuffle<br>color channels | Yes | No |
| **GaussNoise** | Add Gaussian<br>noise | Yes | No |
| **MotionBlur** | Apply motion<br>blur | Yes | No |
| **Blur** | Apply image<br>blurring | Yes | No |
| **OpticalDistortion** | Apply optical<br>warping<br>distortion | Yes | No |
| **GridDistortion** | Apply grid-based<br>geometric<br>distortion | Yes | No |
| **CoarseDropout** | Randomly<br>remove<br>rectangular<br>regions<br>(simulating<br>occlusion) | Yes | No |
| **Sharpen** | Apply sharpening<br>filter | Yes | No |

Case 1 includes anatomically conservative augmentations intended to preserve the integrity of retinal fundus images, while Case 2 consists of general-purpose transformations commonly used in image classification.
Abbreviations: CLAHE, contrast-limited adaptive histogram equalization; RGB, red-green-blue color space; Dropout, random removal of image regions to simulate occlusion.

### Model Architecture and Training

We developed a deep learning model to predict the presence of METS based on fundus images. As shown in Figure 1, fundus images were input into a shared convolutional backbone for feature extraction. We employed a multi-task learning strategy, where the main task was binary classification of METS status, and the auxiliary task was regression of a related clinical parameter such as AC. During training, both tasks were optimized simultaneously, but only the binary classification output for METS was used for the final prediction. Loss functions included binary cross-entropy (BCE) for classification and mean squared error (MSE) for regression, with a loss weighting of 0.8:0.2.

**Fig 1.**
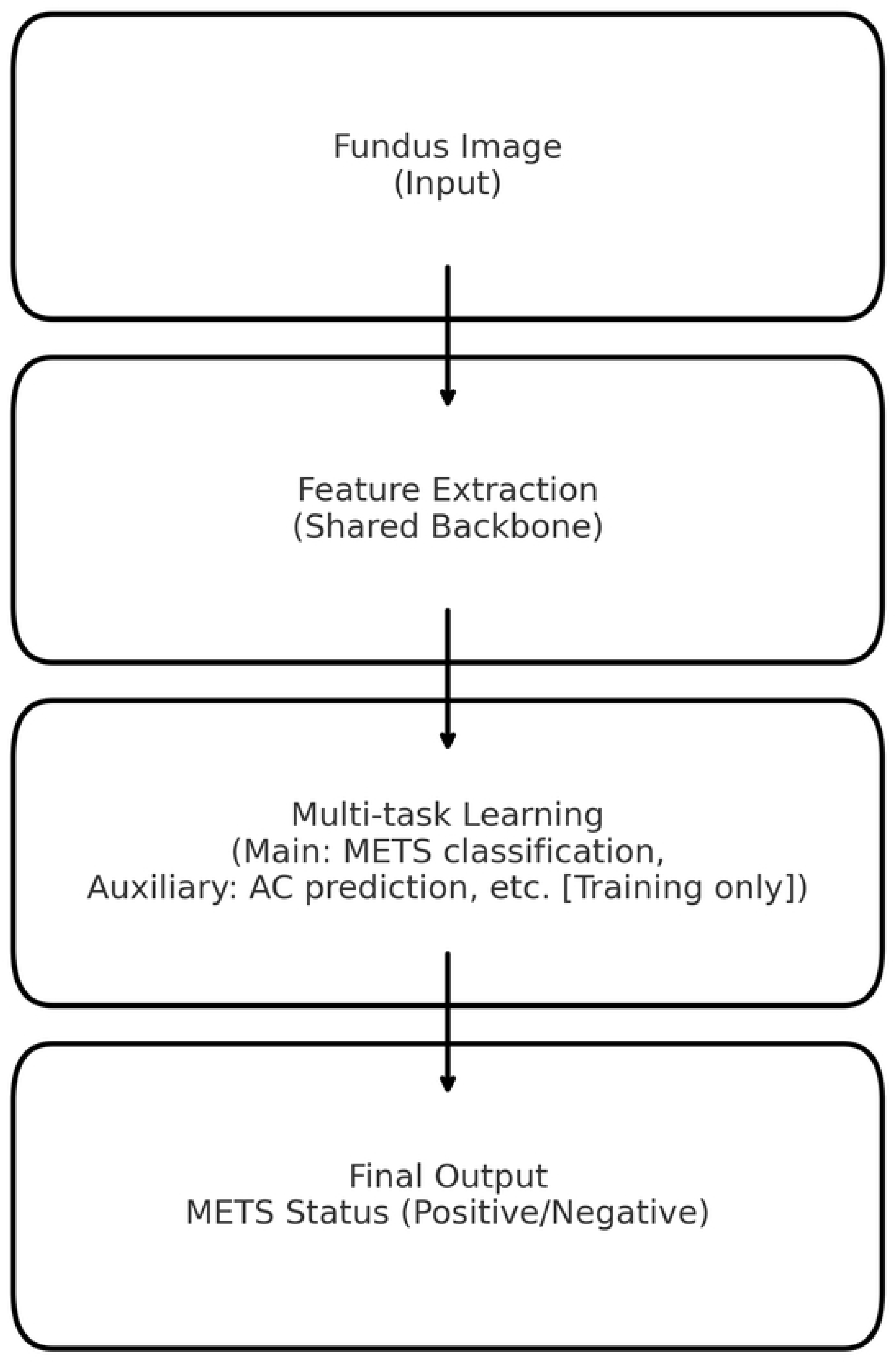
**Overview of the multi-task learning framework for noninvasive prediction of metabolic syndrome (METS) using fundus images.** Fundus images were used as input to a convolutional neural network (shared backbone) for feature extraction. The model employed a multi-task learning strategy, with the main task being binary classification of metabolic syndrome (METS) status, and the auxiliary task being regression of clinical parameters such as abdominal circumference (AC) during training. Final output was the binary classification of METS presence (positive/negative).

Initially, ResNet-50 was used as the backbone, and for final performance evaluation, ConvNeXt-Base, SE-ResNeXt-50, and Swin Transformer V2 Base architectures were also employed. Swin Transformer V2 Base used global average pooling. In contrast, ConvNeXt-Base and SE-ResNeXt-50 employed Generalized Mean (GeM) pooling in place of conventional global average pooling to improve spatial feature aggregation and enhance generalization. The dropout rate was increased to 0.5 before the final classification layers in all models.

### Model Training and Evaluation

Each model was trained using stratified five-fold cross-validation. To assess the impact of different auxiliary variables and augmentation strategies, performance was recorded across folds using the training dataset. Final model performance was evaluated using the independent 500-case test dataset provided by the competition organizers. At the time of model submission, ground truth labels for this test dataset were not disclosed to participants. After the conclusion of the competition, these labels were publicly released by the organizers. All test accuracy and AUC values reported in this study were recalculated by the authors using the released labels.

Three advanced architectures—ConvNeXt-Base, SE-ResNeXt-50, and Swin Transformer V2 Base—were trained using the optimal configuration (Case 1 augmentation + AC auxiliary loss). Their predictions were ensembled by averaging outputs from five cross-validated models for each architecture, followed by average fusion across the three architectures.

TTA was applied only to the final ensemble model. Each test image was evaluated in three forms: original, rotated, and vertically flipped. Final predictions were obtained by averaging these three outputs. Performance metrics reported in the Results section are based solely on the 500-case independent test dataset. To ensure reproducibility, all models were retrained in a standardized software environment with consistent preprocessing and fixed library versions.

### Implementation Environment

All models were implemented in Python 3.12 using PyTorch 2.4.1 and trained on a workstation equipped with an NVIDIA RTX 3090 GPU. Image preprocessing and augmentation were conducted using the Albumentations 1.4.7 library. Model prototyping and configuration management were supported using EyeAIRT (Sensor to AI, Inc.).

## Results

### 3.1 Comparison of Auxiliary Loss Variables and Validation Strategy

We investigated the effect of different auxiliary loss configurations on validation performance in multi-task learning models using ConvNeXt-Base, SE-ResNeXt-50 and Swin Transformer V2 Base architectures. Each model was trained to classify METS status either without auxiliary loss (Main only), or with various clinical parameters as auxiliary regression targets. The evaluated configurations included: AC only; AC combined with SBP and DBP; and all four variables—AC, SBP, DBP, TG, and FPG. As shown in Table 2, the inclusion of AC consistently improved validation accuracy and AUC in both architectures. While adding further variables occasionally led to marginal gains, the overall improvement compared to the AC-only setting was limited and inconsistent.

**Table 2.** Validation accuracy and area under the receiver operating characteristic curve (AUC) for different auxiliary loss configurations in ConvNeXt-Base, SE-ResNeXt-50 and Swin Transformer V2 Base.

| **Model**<br>**Architecture** | **Auxiliary Loss**<br>**Configuration** | **Validation**<br>**Accuracy** | **Validation AUC** |
| --- | --- | --- | --- |
| **ConvNeXt-Base** | **METS**<br>**classification**<br>**only (single-**<br>**task model)** | 0.64828 | 0.70083 |
| **ConvNeXt-Base** | **METS + AC** | 0.66290 | 0.71341 |
| **ConvNeXt-Base** | **METS + AC +**<br>**SBP + DBP** | 0.65308 | 0.71090 |
| **ConvNeXt-Base** | **METS + AC +**<br>**SBP + DBP +**<br>**TG + FPG** | 0.65873 | 0.71242 |
| **SE-ResNeXt-50** | **METS**<br>**classification**<br>**only (single-**<br>**task model)** | 0.65601 | 0.70673 |
| **SE-ResNeXt-50** | **METS + AC** | 0.65559 | 0.70344 |
| **SE-ResNeXt-50** | **METS + AC +**<br>**SBP + DBP** | 0.65078 | 0.69909 |
| **SE-ResNeXt-50** | **METS + AC +**<br>**SBP + DBP +**<br>**TG + FPG** | 0.65392 | 0.69889 |
| **Swin**<br>**Transformer**<br>**V2 Base** | **METS**<br>**classification**<br>**only (single-**<br>**task model)** | 0.64911 | 0.69341 |
| **Swin**<br>**Transformer**<br>**V2 Base** | **METS + AC** | 0.65350 | 0.70514 |
| **Swin**<br>**Transformer**<br>**V2 Base** | **METS + AC +**<br>**SBP + DBP** | 0.65141 | 0.70788 |
| **Swin**<br>**Transformer**<br>**V2 Base** | **METS + AC +**<br>**SBP + DBP +**<br>**TG + FPG** | 0.65622 | 0.71051 |

Validation accuracy and area under the receiver operating characteristic curve (AUC) for multi-task learning models trained under various auxiliary loss configurations. ConvNeXt-Base, SE-ResNeXt-50 and Swin Transformer V2 Base were evaluated using four configurations: no auxiliary loss (“Main only”), abdominal circumference (AC) only, AC + systolic and diastolic blood pressure (SBP, DBP), and AC + SBP + DBP + triglycerides (TG) + fasting plasma glucose (FPG). Abbreviations: METS, metabolic syndrome; AC, abdominal circumference; SBP, systolic blood pressure; DBP, diastolic blood
pressure; TG, triglycerides; FPG, fasting plasma glucose. AUC, area under the receiver operating characteristic curve

Based on these findings, the AC-only configuration was selected for all subsequent training and evaluation. This approach balances predictive performance with model simplicity and interpretability, and was consistently effective across all evaluated architectures.

### 3.2 Comparison of Data Augmentation Strategies

We compared the effects of two different data augmentation strategies: Case 1 (fundus-specific augmentation) and Case 2 (general-purpose augmentation). As shown in Figure 3, the learning curves for training and validation accuracy over 30 epochs revealed that Case 1 achieved more stable convergence and higher validation performance compared to Case 2. While Figure 3 illustrates the training dynamics, a quantitative comparison of validation performance between the two strategies is provided below. As summarized in Table 3, the validation accuracy of the Case 1 models was consistently higher than that of the Case 2 models across different architectures. Specifically, Case 1 achieved a higher average validation accuracy (0.66290) compared to Case 2 (0.63762), supporting its effectiveness in fundus-specific model development.

**Fig 2.**
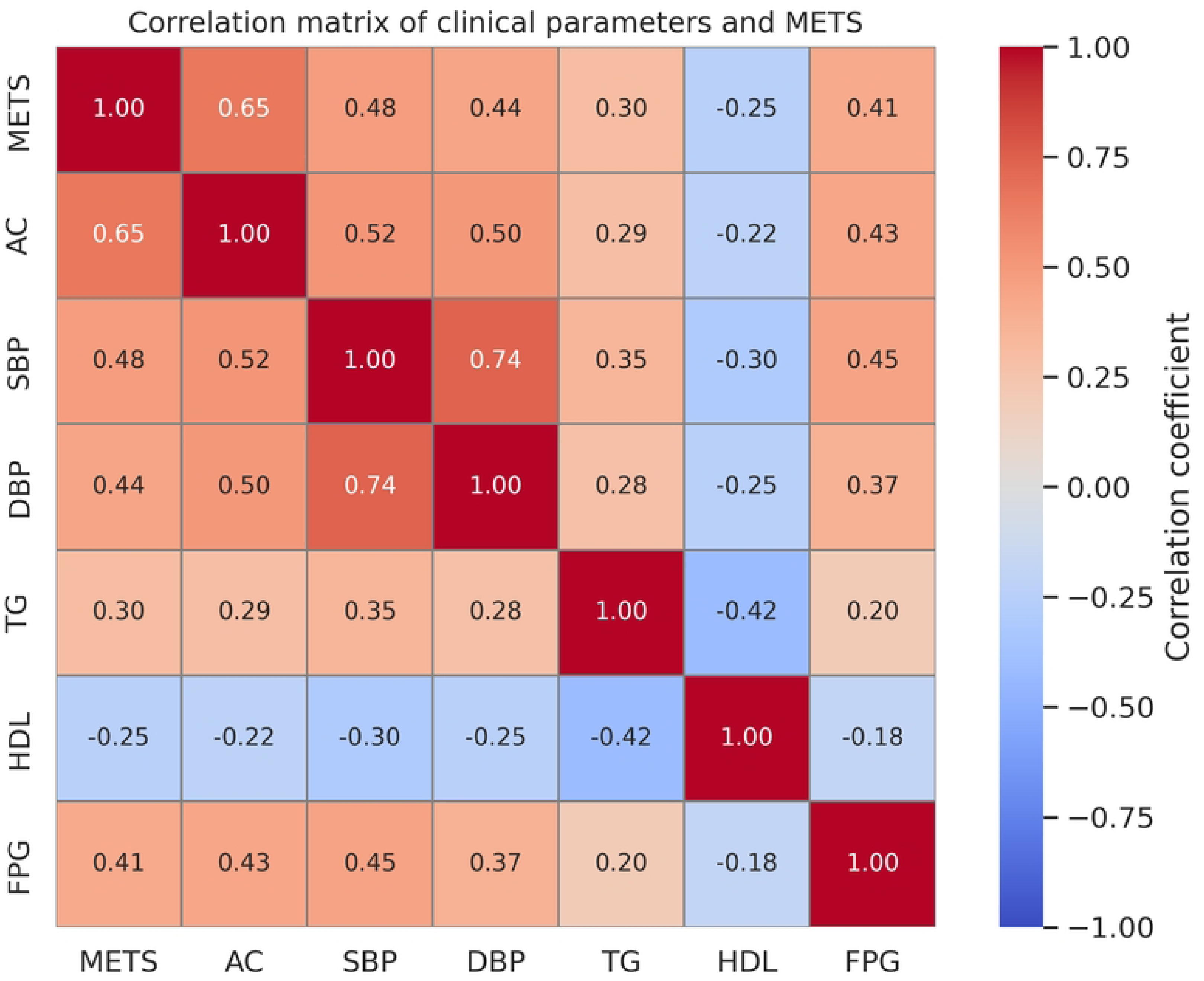
**Correlation matrix between metabolic syndrome (METS) status and clinical indicators in the training dataset.** Abdominal circumference (AC) showed the strongest positive correlation with the presence of metabolic syndrome (METS), followed by systolic blood pressure (SBP) and diastolic blood pressure (DBP). The results guided the selection of auxiliary variables for the multi-task learning framework. Abbreviations: METS, metabolic syndrome; AC, abdominal circumference; SBP, systolic blood pressure; DBP, diastolic blood pressure; TG, triglycerides; HDL, high-density lipoprotein cholesterol; FPG, fasting plasma glucose.

**Fig 3A.**
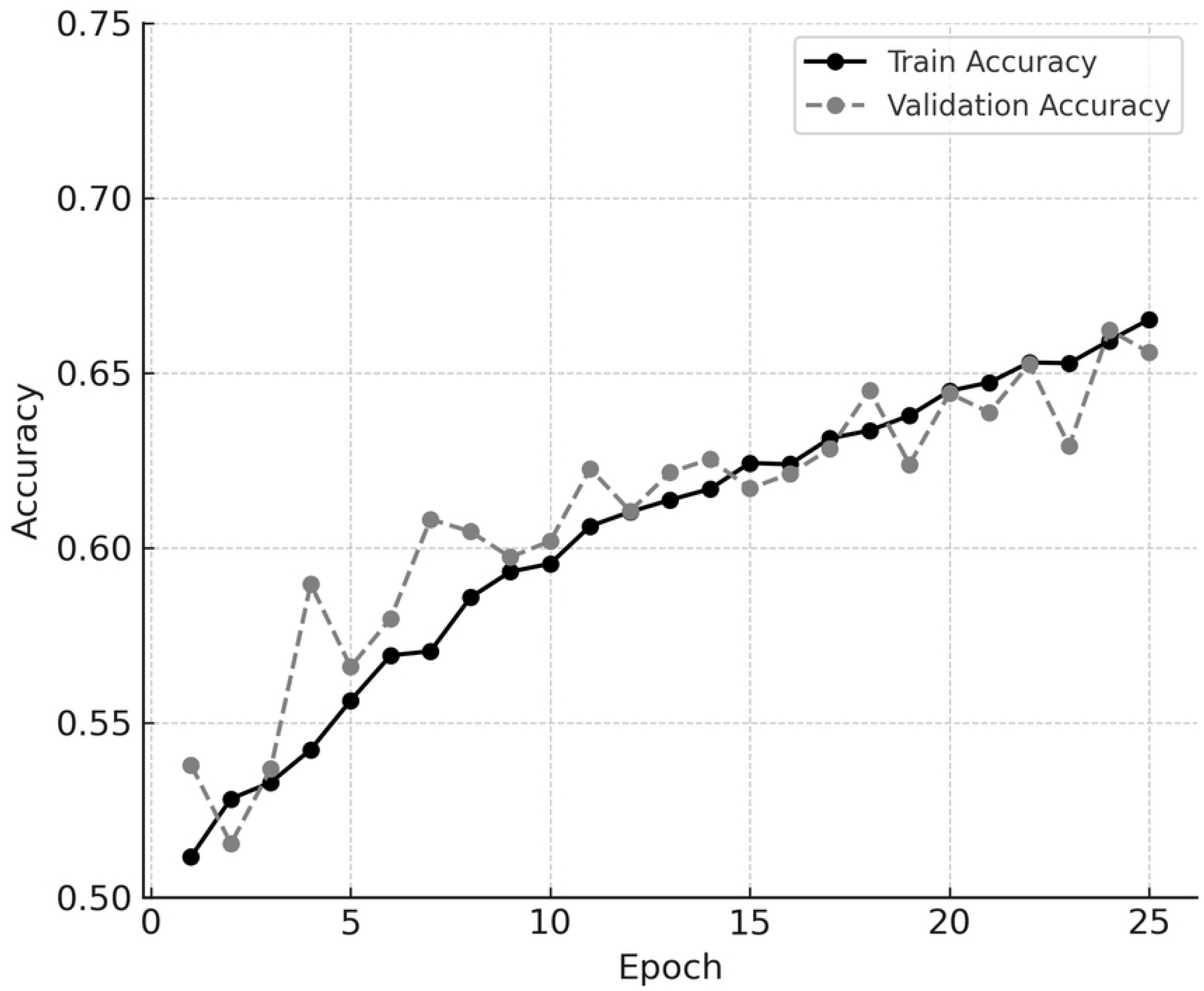
**Training and validation accuracy curves for Case 1** Learning curves for Case 1 using fundus-specific data augmentation. Training accuracy and validation accuracy both increase steadily over the course of 25 epochs, with only a small gap between them. This indicates that the model trained with fundus-specific augmentation achieved stable convergence and generalization without signs of overfitting.

**Fig 3B.**
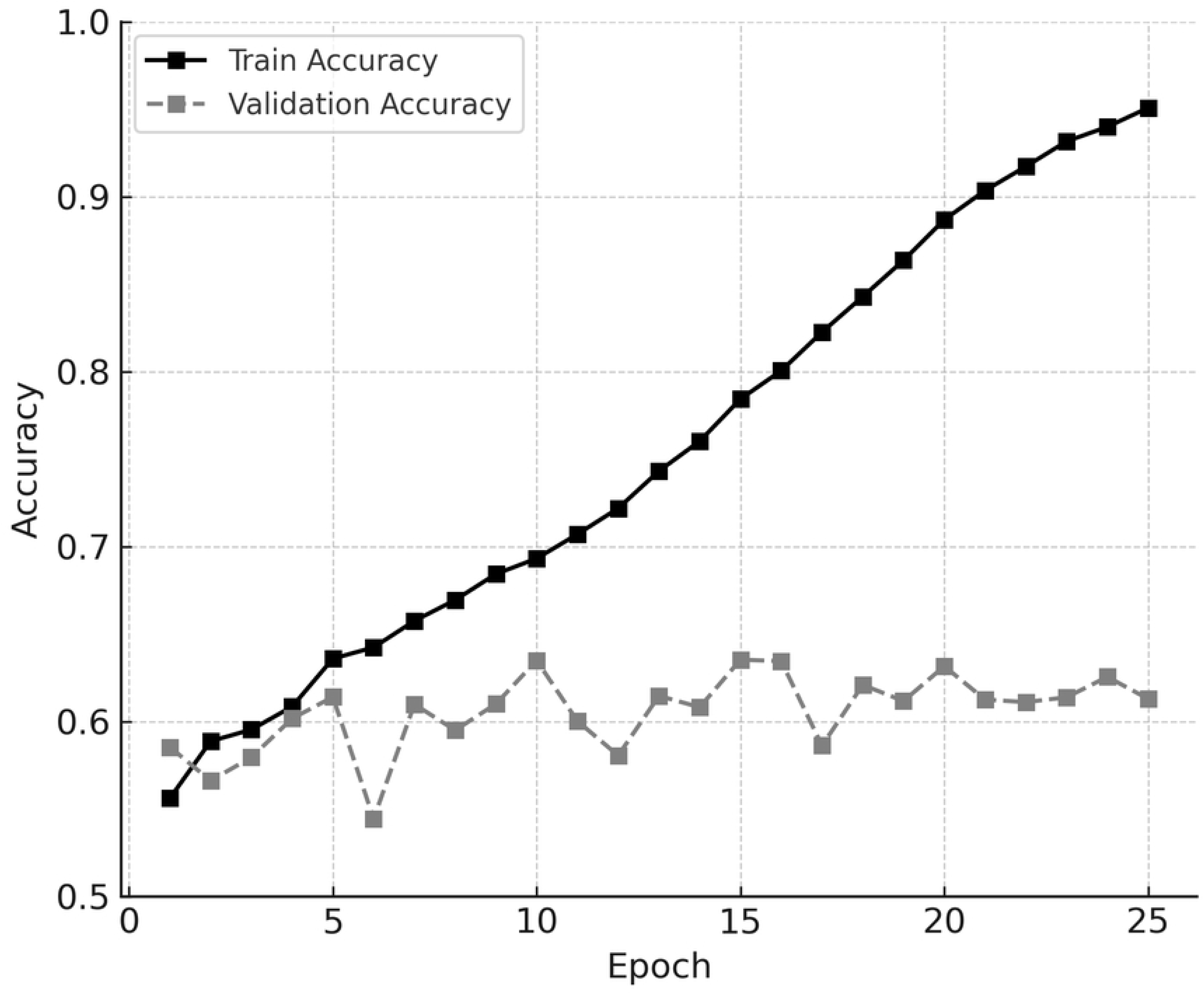
**Training and validation accuracy curves for Case 2** Learning curves for Case 2 using general-purpose data augmentation. While training accuracy increases continuously, validation accuracy plateaus early and then gradually declines. This pattern suggests that the model is overfitting the training data and fails to generalize effectively to unseen data.

**Table 3.** Validation accuracy and training stability for different augmentation strategies.

| **Augmentation**<br>**Strategy** | **Mean Validation**<br>**Accuracy** | **Mean Validation**<br>**AUC** | **Mean Epoch of**<br>**Best Validation**<br>**Loss** |
| --- | --- | --- | --- |
| **Case 1 (Fundus-**<br>**specific)** | 0.66290 | 0.71341 | 22.6 |
| **Case 2**<br>**(General-**<br>**purpose)** | 0.63762 | 0.68665 | 6.0 |

Comparison of two data augmentation strategies based on average validation accuracy and training stability across five-fold cross-validation. Case 1 incorporated anatomically conservative transformations tailored for fundus images, while Case 2 used general-purpose augmentations. Note: Test data were not used for this comparison; only validation performance during model development was considered. Abbreviations: AUC, area under the receiver operating characteristic curve

### 3.3 Final Model Performance with Ensemble and TTA

To evaluate the final classification performance of our system, we tested three advanced architectures—ConvNeXt-Base, SE-ResNeXt-50 and Swin Transformer V2 Base —on the independent test dataset comprising 500 cases. All models were trained under the optimal configuration using fundus-specific augmentation (Case 1) and AC as the auxiliary task. As shown in Table 4, the individual models achieved test accuracies of 0.66400 (ConvNeXt-Base), 0.66000 (SE-ResNeXt-50), and 0.66600 (Swin Transformer V2 Base), with corresponding AUCs of 0.72136, 0.70139, and 0.71712, respectively.

**Table 4.**
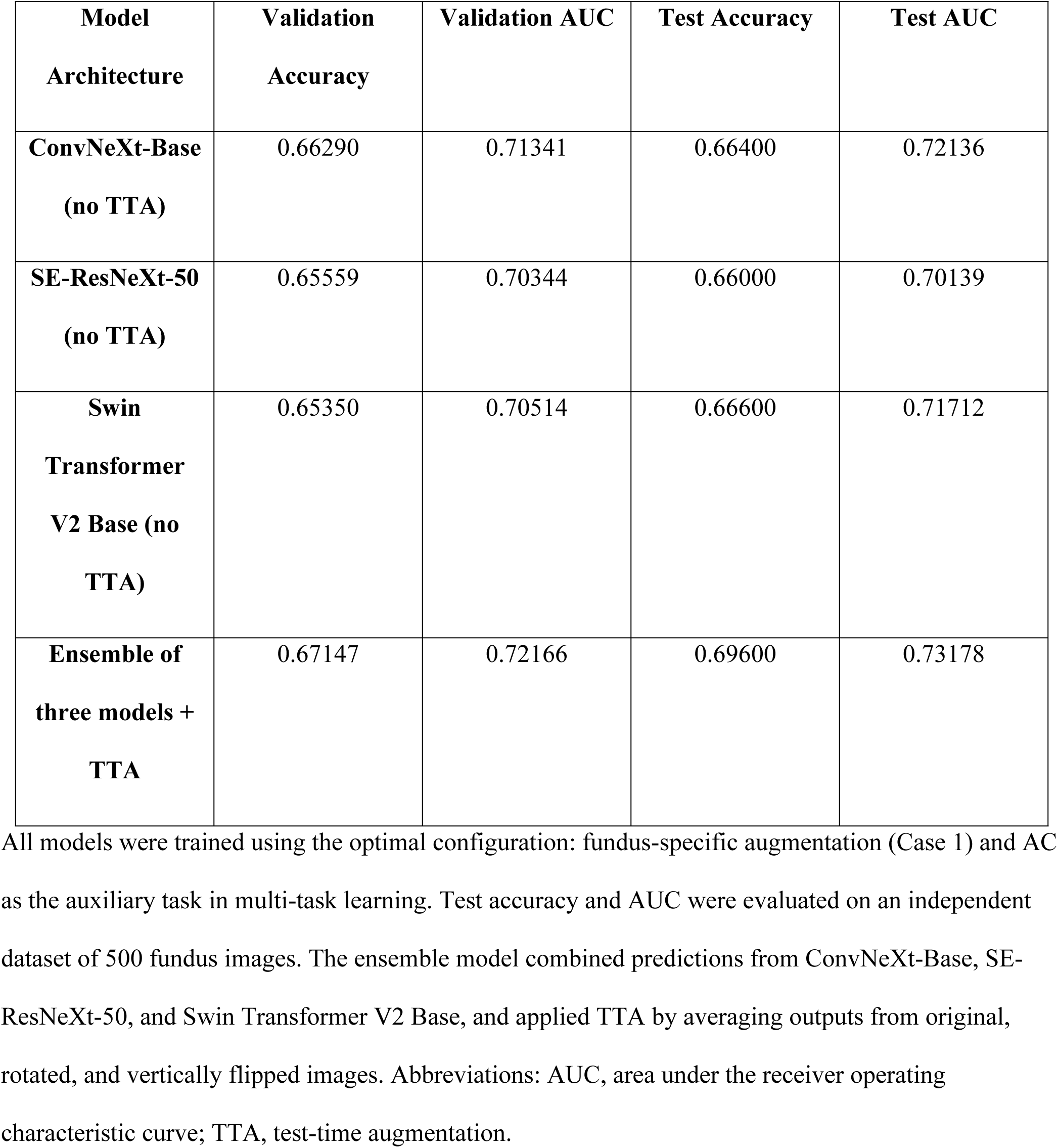
Final test performance of individual models and the ensemble model.

| **Model**<br>**Architecture** | **Validation**<br>**Accuracy** | **Validation AUC** | **Test Accuracy** | **Test AUC** |
| --- | --- | --- | --- | --- |
| **ConvNeXt-Base**<br>**(no TTA)** | 0.66290 | 0.71341 | 0.66400 | 0.72136 |
| **SE-ResNeXt-50**<br>**(no TTA)** | 0.65559 | 0.70344 | 0.66000 | 0.70139 |
| **Swin**<br>**Transformer**<br>**V2 Base (no**<br>**TTA)** | 0.65350 | 0.70514 | 0.66600 | 0.71712 |
| **Ensemble of**<br>**three models +**<br>**TTA** | 0.67147 | 0.72166 | 0.69600 | 0.73178 |

All models were trained using the optimal configuration: fundus-specific augmentation (Case 1) and AC as the auxiliary task in multi-task learning. Test accuracy and AUC were evaluated on an independent dataset of 500 fundus images. The ensemble model combined predictions from ConvNeXt-Base, SE- ResNeXt-50, and Swin Transformer V2 Base, and applied TTA by averaging outputs from original, rotated, and vertically flipped images. Abbreviations: AUC, area under the receiver operating characteristic curve; TTA, test-time augmentation.

Subsequently, we constructed an ensemble model by averaging the predictions from all three architectures and applying TTA. Each test image was evaluated in three forms (original, rotated, vertically flipped), and the final prediction was determined by averaging the outputs. The ensemble model with TTA achieved a test accuracy of 0.69600 and an AUC of 0.73178, outperforming all individual models. While this configuration yielded the highest performance, the improvement may partly reflect the complementary nature of the integrated architectures rather than TTA alone. These findings support the value of ensemble learning in fundus image-based classification tasks.

## Discussion

In this study, we developed a multi-task deep learning model to predict METS from retinal fundus images obtained from Japanese health checkup participants. The final model achieved a test accuracy of 0.696 through the integration of AC as an auxiliary task, fundus-specific data augmentation, and ensemble learning with TTA. These results demonstrate the feasibility of non-invasive, image-based METS screening using limited but well-curated retinal datasets.

The use of AC as an auxiliary variable in a multi-task learning framework was found to be the most effective among the clinical indicators tested. This finding is based on validation accuracy observed during model development, where the configuration using AC alone as an auxiliary loss yielded the best performance. This highlights the importance of selecting auxiliary tasks that are semantically and visually aligned with the primary classification target, thereby minimizing gradient interference during training [4]. Since AC reflects visceral fat accumulation—a central factor in METS—it may be indirectly manifested in the structure and appearance of the retinal vasculature. Epidemiological studies have also reported significant associations between retinal microvascular signs and METS components, including AC [3].

Our results also emphasize the benefits of fundus-specific data augmentation strategies. Based on validation accuracy observed during cross-validation, Case 1 augmentation—which included anatomically conservative transformations tailored to retinal fundus images—consistently outperformed the general-purpose Case 2 strategy. Case 1 led to more stable training and higher mean accuracy, while Case 2 resulted in early convergence and reduced performance. This suggests that domain-aware preprocessing plays a critical role in improving deep learning performance, particularly in medical image analysis where anatomical orientation is clinically meaningful.

The final model leveraged three advanced neural network architectures—ConvNeXt-Base, SE-ResNeXt-50, and Swin Transformer V2 Base —and demonstrated improved predictive performance through ensemble learning and TTA. Such strategies are known to enhance robustness and consistency across medical imaging tasks [5], and this was validated in our study using a 500-case independent test set.

In addition to accuracy, the AUC was also evaluated. The final ensemble model achieved an AUC of 0.73178, while individual models recorded AUCs of 0.72136 (ConvNeXt-Base), 0.70139 (SE-ResNeXt-50), and 0.71712 (Swin Transformer V2 Base), respectively. These values indicate strong discriminatory performance. Notably, Cheung et al. reported an AUC of 0.744 using over 13,000 retinal images [7]; our model achieved comparable performance using only 5,000 cases, highlighting the effectiveness of our task-specific optimization and augmentation strategies even in smaller datasets.

In the early phase of model development, we considered incorporating multiple clinical parameters related to metabolic syndrome, such as obesity, blood pressure, HbA1c, and lipid profiles, into the auxiliary tasks of multi-task learning. However, based on preliminary correlation analysis within the training dataset, we found that AC exhibited the strongest association with METS status among the candidates. Given the time constraints during the competition phase, we initially adopted a focused strategy by selecting AC as the sole auxiliary task. Subsequent analyses conducted for the purpose of formal publication reaffirmed that the use of AC alone provided the most stable and highest predictive performance for METS. These findings suggest that in multi-task learning, careful selection of auxiliary tasks that are strongly and directly linked to the main prediction target is critical for optimizing model performance. Rather than indiscriminately including multiple sub-tasks, strategic focusing based on clinical and statistical relevance can significantly contribute to achieving robust results. Furthermore, the strategic selection of AC as an auxiliary task based on preliminary correlation analysis is a novel aspect of this study. While multi-task learning has been explored in other medical imaging domains, its application to fundus-based metabolic risk prediction with systematic auxiliary variable selection has been scarcely reported. Our findings highlight the potential of auxiliary loss design in enhancing the predictive performance of fundus image-based AI models.

Recent advances in “oculomics”—the study of systemic health markers through ocular imaging— have further underscored the potential of the retina as a biomarker-rich window into overall physiology. Poplin et al. [6] reported that deep learning could predict cardiovascular risk factors such as age, blood pressure, and smoking status from fundus images with AUCs ranging from 0.70 to 0.82, depending on the variable. Similarly, Rim et al. demonstrated that AI models could infer kidney function markers such as estimated glomerular filtration rate (eGFR) from fundus photographs [9]. Other studies have shown that fundus-based AI can accurately estimate hemoglobin levels [10], biological age [11], and even cognitive decline [12], highlighting the retina’s unique role in reflecting multi-organ health. Our study contributes to this expanding body of oculomic research by focusing on METS prediction and incorporating multi-task learning to enhance clinical interpretability. In contrast to previous single-task approaches, our model’s auxiliary loss design guided by clinical correlation strengthens its connection to physiologically meaningful features.

This study has several limitations. First, the dataset provided for training consisted solely of male participants. While this reduced variability and improved model stability, it limits the generalizability of the results to female populations. Future work should explore sex-stratified model development and validation. Second, the auxiliary loss was designed using only a limited number of clinical variables. Incorporating additional predictors—such as HbA1c, insulin resistance markers, or lifestyle factors like smoking and physical activity—may further improve performance. Third, all fundus images used in this study were captured using Canon retinal cameras, as confirmed via image metadata. While this ensured consistency in image quality, the generalizability of the model to images captured by different devices or under varied conditions remains to be evaluated through external validation.

## Conclusion

This study presents a reproducible and accurate deep learning framework for predicting METS from retinal fundus images in a Japanese health checkup cohort. Through the integration of multi-task learning, fundus-specific data augmentation, and ensemble prediction with TTA, our model achieved clinically meaningful performance while using relatively limited training data. These findings support the potential for scalable, non-invasive AI-based screening tools for systemic diseases using retinal imaging, particularly in preventive medicine and public health settings. In addition, the strategic design of the auxiliary loss function, guided by preliminary clinical correlation analysis, represents a novel contribution to fundus-based AI research.

## Data Availability

The fundus image dataset analyzed during the current study is not publicly available due to privacy and ethical restrictions. However, anonymized data may be made available from the corresponding author upon reasonable request and with appropriate institutional approval.

## Acknowledgments

The clinical information and fundus photographs used in this study were provided by the Japanese Society of Artificial Intelligence in Ophthalmology and the National Institute of Informatics, and were managed by the Japan Ocular Imaging Registry website (http://joir.jp/).

## Competing Interests

K.N., T.I., and S.W. have received consulting fees from SAI Inc. Y.F. is the President of SAI Inc., the company that provides the Eye AIRT technology used in this study. This does not alter our adherence to PLOS ONE policies on sharing data and materials.

## Notes

### Competing Interest Statement

I have read the journal's policy and the authors of this manuscript have the following competing interests: K.N., K.I., and Y.W. have received consulting fees from SAI Inc. Y.F. is the President of SAI Inc., the provider of the Eye AIRT technology used in this study. These relationships are unrelated to the submitted work.

### Funding Statement

The author(s) received no specific funding for this work.

### Author Declarations

This study used existing, anonymized data provided for research purposes through the AI model development competition organized by the Japanese Society of Ophthalmic AI. According to the competition’s data usage policy, ethical review and approval were not required.

